# Discrepancies in Interpretation of Japanese Radiology Report Expressions Between Radiologists and Referring Physicians

**DOI:** 10.64898/2026.10.19.26364709

**Authors:** Takeru Yamaguchi, Hidetoshi Matsuo, Mizuho Nishio, Junko Inoue Inukai, Takaaki Matsunaga, Yasuyuki Kojita, Keitaro Sofue, Takamichi Murakami

**Affiliations:** Department of Radiology, Kobe University Graduate School of Medicine, 7-5-2, Kusunokicho, Chuo-ku, Kobe, Hyogo, 650-0017, Japan

**Keywords:** radiology report, interpretation, communication, radiologists, referring physicians

## Abstract

**Objectives:** To evaluate discrepancies between radiologists and referring physicians in the interpretation of diagnostic probabilities conveyed by common Japanese expressions used in radiology reports.

**Materials and Methods:** Fifteen expressions commonly used to convey diagnostic probabilities in radiology reports were selected by five attending radiologists. An anonymous online survey was distributed to 26 radiologists and 33 referring physicians at a single institution. Participants assigned a diagnostic probability (from 0% to 100% in 10% increments) to each expression. Differences in median probabilities between radiologists and referring physicians were analyzed using the Mann–Whitney U test. Variability in interpretation was assessed using interquartile range (IQR), with IQR ≥ 30% defined as substantial variability.

**Results:** All radiologists (26/26) and 97% of referring physicians (32/33) completed the survey. Seven of the 15 expressions showed statistically significant differences in interpretation between radiologists and referring physicians: “c/w” (80% vs. 60%; *p* = 0.031), the Japanese equivalents of “present” (100% vs. 100%; *p* = 0.024), “suspect of” (80% vs. 70%; *p* = 0.007), “possibly” (50% vs. 70%; *p* < 0.001), “unlikely” (10% vs. 10%; *p* = 0.022), “cannot be ruled out” (25% vs. 40%; *p* < 0.001), and “not inconsistent with” (80% vs. 90%; *p* = 0.008). Substantial variability in interpretation (IQR ≥ 30%) was observed for four expressions: “cannot be ruled out” (IQR, 10%–40%), “s/o” (IQR, 60%–90%), “c/w” (IQR, 50%–90%), and “not inconsistent with” (IQR, 70%–100%).

**Conclusion:** This study revealed that nearly half of the Japanese expressions commonly used to convey diagnostic probabilities were interpreted differently by radiologists and referring physicians. These discrepancies highlight the need for a standardized lexicon to improve clarity and consistency in radiology reporting.

**Compliance with Reporting Guidelines:** This study was conducted in accordance with the STROBE guidelines for observational studies.

## Introduction

One of the most critical roles of radiologists in clinical practice is to generate radiology reports that convey the meaning and clinical relevance of imaging findings.^[1]^ Clear and precise reporting is critical for effective communication with the referring physicians.^[2]^ Accurately conveying diagnostic certainty is one of the most crucial elements of high-quality radiology reports, as it enables referring physicians to make informed decisions regarding patient management.^[1,3]^ These decisions, including further diagnostic procedures, surgical interventions, and other treatments, can significantly influence patient outcomes.

To convey levels of diagnostic certainty, radiologists use various expressions (e.g., “diagnostic of,” “likely,” “possibly”). Previous English studies have evaluated the diagnostic probabilities associated with these terms.^[4–9]^ These studies have consistently demonstrated a low concordance among radiologists and between radiologists and referring physicians in the interpretation of these expressions. These findings underscore the importance of standardized terminology to convey probability and the use of clear, unambiguous language in radiology reporting.^[10]^ However, our literature review found no prior studies that evaluated the concordance of diagnostic probability interpretations for radiology report phrasing in languages other than English. Investigating radiology report phrasing in non-English languages is important because language and cultural differences may influence how diagnostic certainty is conveyed and interpreted, potentially impacting clinical decision-making and patient outcomes.

Thus, the purpose of this study was to evaluate differences and variability in the interpretation of diagnostic probability for commonly used Japanese radiology report expressions between radiologists and referring physicians.

## Materials and methods

This institutional review board-approved study was conducted at a large academic hospital in Japan with over 900 beds and a radiology department that generates more than 69,000 radiology reports annually for computed tomography (CT) and magnetic resonance (MR) scans. The requirement for informed consent from patients was waived.

### Survey Design and Content

Five attending radiologists discussed selecting 15 commonly used Japanese expressions that describe probability, such as “o utagau (suspected)” (Table 1). The influence of these terms on the perceived likelihood of “true metastatic disease” was subsequently evaluated by requesting both radiologists and referring physicians to assign a statistical probability of “true metastatic disease” based on the specific terminology used.^[7]^ An anonymous online survey was administered (Supplementary Material). Respondents were asked to indicate their department (internal medicine or surgical specialties) and years of clinical experience and to select the degree of diagnostic probability for each of the 15 phrases from the following 11 options: 0%, 10%, 20%, 30%, 40%, 50%, 60%, 70%, 80%, 90%, and 100%. The 15 expressions were presented in a randomized order in the questionnaire.

**Table 1.** Japanese Diagnostic Probability Expressions Evaluated in the Study.

| Japanese Expression | English Translation |
| --- | --- |
| s/o X | abbreviation of "suspicious of X" |
| c/w X | abbreviation of "consistent with X" |
| X ari | X is present |
| X utagai | suspicious of X |
| X o utagau | X is suspected |
| X to kangaeru | X is considered |
| X wa hiteiteki | negative for X |
| X o mitomeru | X is observed |
| X no kanosei ari | possibly X / possibility of X exists |
| X wa kangae nikui | X is unlikely / hard to consider X |
| X no kanosei wa hikui | possibility of X is low |
| X wa shiteki dekinai | X cannot be identified |
| X o utagau shoken nashi | no findings suggestive of X |
| X o hitei dekinai | X cannot be ruled out |
| X to shite mujun shinai | not inconsistent with X |

X indicates the suspected lesion or diagnosis.

### Participants

All radiologists at our institution with 5–30 years of clinical experience who routinely interpret CT and MR examinations were invited to participate in the survey (*n* = 26).

Referring physicians were selected from our institution’s physician registry to ensure a balanced distribution in terms of specialty and years of clinical experience (5–30 years). No more than four physicians were selected from a single department. Assuming an anticipated response rate of 80% or higher, a total of 33 referring physicians were solicited to ensure that the number of respondents would be at least equal to or greater than that of the radiologists. Physicians were solicited via phone call and/or e-mail to participate in the online survey. The specialties of these physicians included urology (*n* = 4), obstetrics and gynecology (*n* = 4), neurology (*n* = 4), neurosurgery (*n* = 3), gastroenterology (*n* = 3), diabetes and endocrinology (*n* = 3), surgery (*n* = 2), cardiology (*n* = 2), respiratory medicine (*n* = 2), general internal medicine (*n* = 2), cardiovascular surgery (*n* = 1), orthopedics (*n* = 1), otolaryngology (*n* = 1), and hematology and oncology (*n* = 1).

### Frequency Analysis of Expressions

To investigate the frequency of each expression used in radiology reports at our institution, we reviewed reports generated during the two-month period preceding the start of the survey (April 1 to May 31, 2025). The frequency of each expression was analyzed separately in the findings section, the impressions section, and the entire report. Expressions not used in the context of conveying diagnostic probability were excluded from the analysis. To avoid double-counting, expressions embedded within longer phrases were counted only as a part of the longer phrase. For example, “o utagau (suspected)” was not counted separately when included in “o utagau shoken nashi (no findings suggestive of).”

### Statistical Analysis

Descriptive statistics, including the median and interquartile range (IQR), were used to summarize the years of clinical experience and the interpretation of probability expressions by the radiologists and referring physicians. The Mann–Whitney U test was used to compare the years of clinical experience and the distribution of estimated probability percentages for each expression between the two groups, as normality was assessed using the D’Agostino– Pearson test and the data were found not to follow a normal distribution. For each expression, responses were treated as continuous variables representing the estimated probability percentages (0%–100%). Variability in interpretation was assessed using the IQR, with IQR ≥ 30% defined as substantial variability.^[9]^ Statistical significance was set at *p* < 0.05. Box plots were generated to visually represent the distribution of responses for each expression in the two groups. All statistical analyses were performed using MedCalc version 23.1.7 (MedCalc Software, Ostend, Belgium).

## Results

All radiologists (26/26, 100%) and 32 of 33 referring physicians (97 %) completed the survey. Among the referring physicians, 17 (53%) specialized in internal medicine and 15 (47%) in surgical specialties. There was no significant difference in the median years of clinical experience between the radiologists and referring physicians (10.5 [IQR, 8–21] vs. 12 [IQR, 9–21]; *p* = 0.51).

The estimated probabilities for each expression, as reported by radiologists and referring physicians, are summarized in Table 2 and visualized as box plots in Figure 1. Among the 15 expressions evaluated, seven showed statistically significant differences in interpretation between radiologists and referring physicians: “c/w” (80% vs. 60%; *p* = 0.031), “ari (present)” (100% vs. 100%; *p* = 0.024), “utagai (suspect of)” (80% vs. 70%; *p* = 0.007), “no kanosei ari (possibly)” (50% vs. 70%; *p* < 0.001), “wa kangae nikui (unlikely)” (10% vs. 10%; *p* = 0.022), “o hitei dekinai (cannot be ruled out)” (25% vs. 40%; *p* < 0.001), and “to shite mujun shinai (not inconsistent with)” (80% vs. 90%; *p* = 0.008) [Figure 1]. No significant differences were observed among the remaining eight expression levels.

**Table 2.**
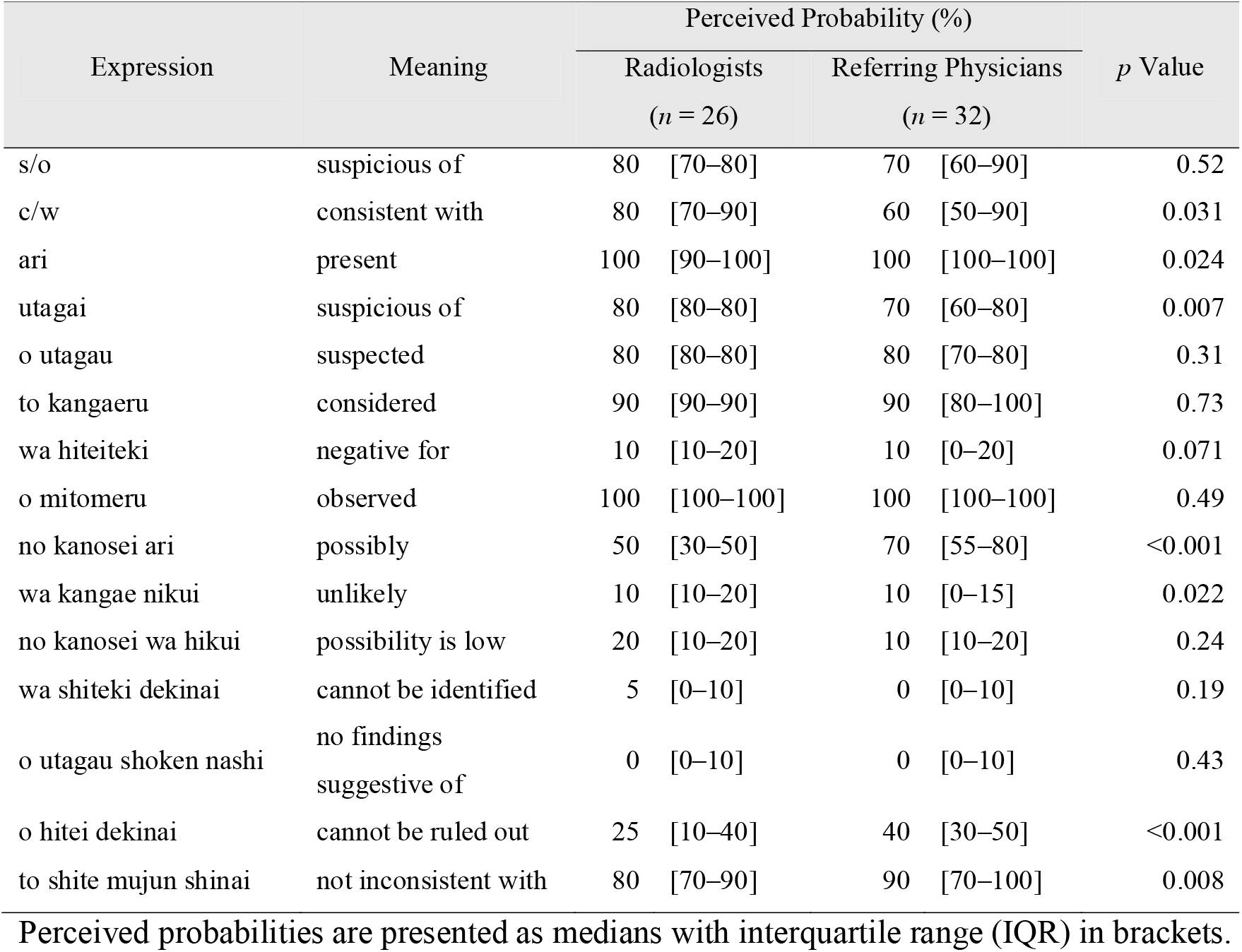
Median and IQR of Perceived Probabilities for Each Expression by Radiologists and Referring Physicians.

**Figure 1.**
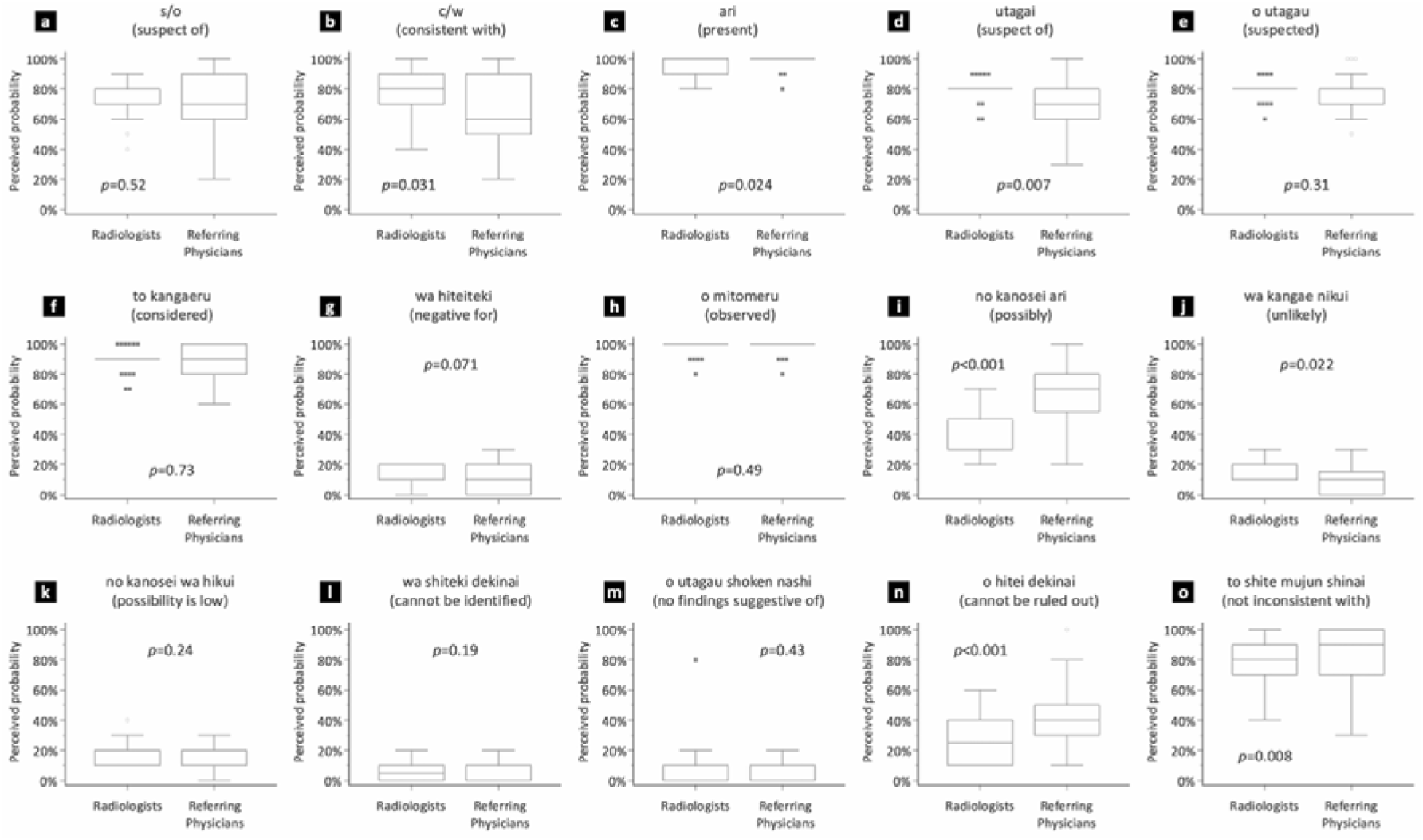
Box plots of perceived probabilities for each expression by radiologists and referring physicians. Each subpanel (a–o) corresponds to a different diagnostic probability expression. Open circles indicate outside values (beyond 1.5 times the interquartile range [IQR] from the quartiles), and filled squares indicate far-out values (beyond 3 times the IQR from the quartiles). Statistical significance was set at *p* < 0.05. Seven expressions showed statistically significant differences between radiologists and referring physicians: (b) “c/w”, (c) “ari (present)”, (d) “utagai (suspect of)”, (i) “no kanosei ari (possibly)”, (j) “wa kangae nikui (unlikely)”, (n) “o hitei dekinai (cannot be ruled out)”, and (o) “to shite mujun shinai (not inconsistent with).”

Among the 15 expressions analyzed, specific phrases exhibited substantial variability in interpretation. Specifically, “o hitei dekinai (cannot be ruled out)” among radiologists (IQR, 10%–40%) and “s/o,” “c/w,” and “to shite mujun shinai (not inconsistent with)” among referring physicians (IQRs, 60%–90%, 50%–90%, and 70%–100%, respectively) showed IQRs ≥ 30%, indicating wide variability in perceived diagnostic probability [Figure 1].

During the 2 months preceding the start of the survey, 11,676 reports were generated for the CT and MR scans. The most frequently used expressions for indicating very high probabilities were “ari (present)” (65%), “utagai (suspicious of)” (46%), “o utagau (suspected)” (28%), and “o mitomeru (observed)” (26%), while “wa shiteki dekinai (cannot be identified)” (34%) was the most common for very low probabilities (Figure 2). In the findings section, the most frequently used expression was “ari (present)” (65%). However, its usage dropped markedly in the impressions section (4.4%), where the most commonly used expression was “utagai (suspicious of)” (20%).

**Figure 2.**
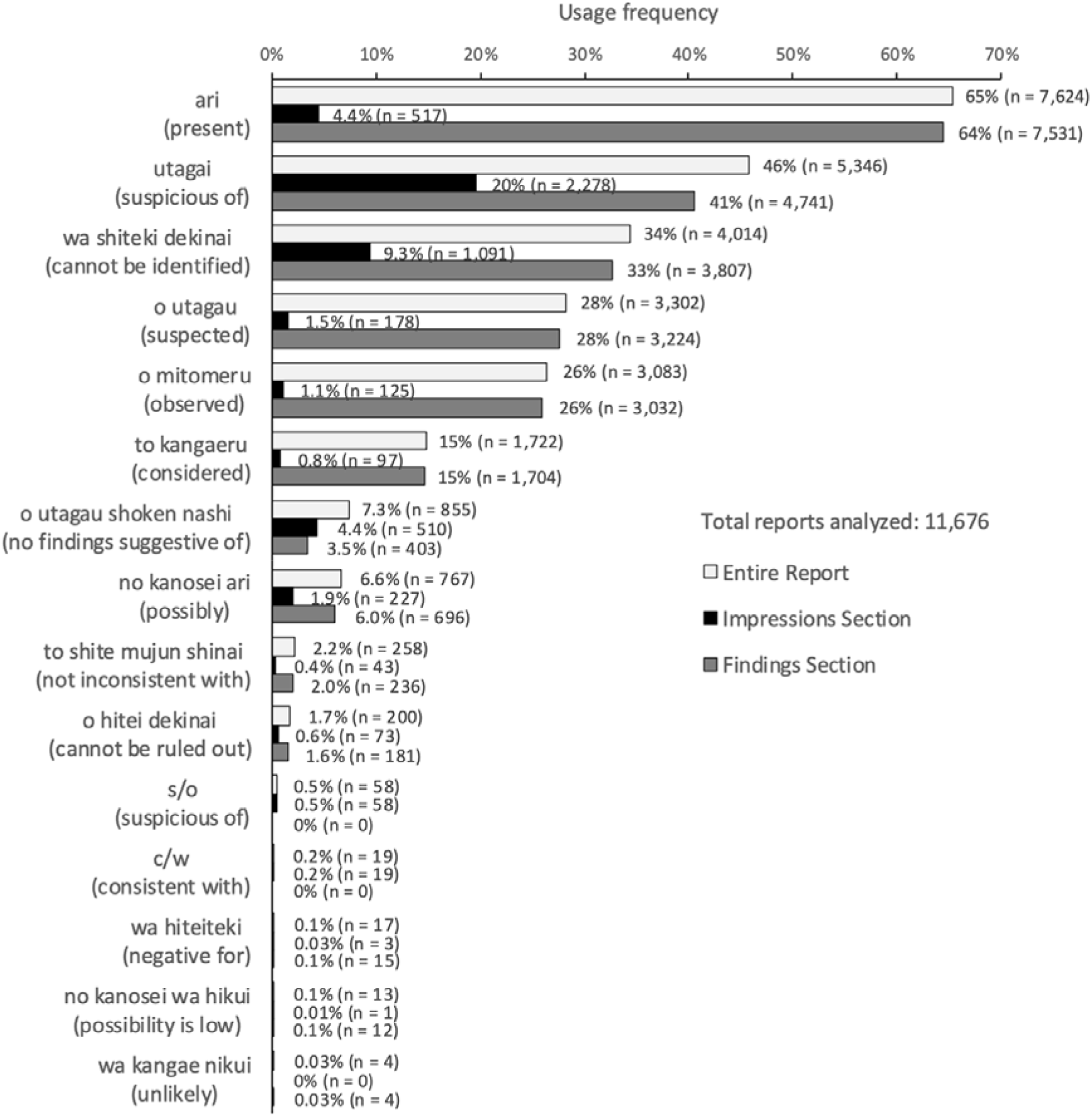
Bar chart showing the usage frequencies of diagnostic probability expressions in radiology reports. Bars represent the percentage of each expression used in the findings section, the impressions section, and the entire report. Percentages and counts are shown next to each bar.

## Discussion

The present study demonstrated that seven of 15 Japanese language expressions describing probability, commonly used in radiology reporting, showed a significant difference in perceived diagnostic probability between radiologists and referring physicians. Four expressions exhibited substantial variability in perceived probability (IQR ≥ 30%) among radiologists and referring physicians.

Previous studies have reported poor concordance both among radiologists and between radiologists and referring physicians in the interpretation of radiology reporting expressions in English-language settings, potentially affecting clinical decisions and patient outcomes.^[4–9]^ In our study, which was conducted in Japanese, significant differences in interpretations were observed for seven phrases commonly used in radiology reporting. Among these, “c/w,” “no kanosei ari (possibly),” and “o hitei dekinai (cannot be ruled out)” showed a difference of 15–20 points in median scores between radiologists and referring physicians, which may have a meaningful impact on clinical decision-making. For example, the expression “no kanosei ari (possibly)” was interpreted with a median probability of 70% by referring physicians, compared to 50% by radiologists. This suggests that referring physicians may perceive a higher likelihood of disease than intended by a radiologist, potentially leading to more aggressive clinical decisions. Such misalignment may result in unnecessary interventions, including biopsy, surgery, or additional imaging associated with radiation exposure, potentially causing patient harm without proportional clinical benefit. To ensure clear and consistent communication, the use of these expressions should be avoided.

Among the seven expressions with significant differences in interpretation between radiologists and referring physicians, variability in perceived probability was considerably high (IQR ≥ 30%) for “c/w” and “to shite mujun shinai (not inconsistent with)” among referring physicians and “o hitei dekinai (cannot be ruled out)” among radiologists, suggesting these expressions have lower consistency in usage and interpretation. In particular, the expression “c/w” demonstrated the most significant variability in interpretation (IQR, 50%–90%). Post-survey informal communication revealed that some physicians were unfamiliar with the meaning of this term, which may have contributed to wide variations in their responses. Avoiding such ambiguous and potentially confusing expressions may help improve the clarity of radiology reports. Based on the range of interpretations shown in the box plots, the expressions indicating very high or very low probabilities were interpreted more consistently than those indicating intermediate probabilities, which is consistent with the findings of a previous study.^[9]^

Frequency analysis revealed that “ari (present)” and “wa shiteki dekinai (cannot be identified)” were the most frequently used expressions to indicate very high and very low possibilities, respectively. Although “ari (present)” was the most commonly used expression in the findings section, its usage declined markedly in the impressions section. A possible explanation is that “ari (present)” is often used to describe frequently observed but clinically insignificant findings (e.g., hepatic cysts), which are less likely to be included in the impressions section. Overall, fewer expressions conveying diagnostic probability were used in the impressions section than in the findings section. This may be partly because radiologists avoid redundant phrasing when making definitive statements. For example, stating simply “liver metastases” in the impressions section rather than “liver metastases are observed” may be preferred among radiologists.

Radiologists must develop effective strategies to communicate diagnostic certainty to improve patient care. One potential solution is to establish a standardized lexicon to express the degree of certainty of radiological diagnoses.^[11]^ Wibmer et al. reported that the implementation of a standardized diagnostic certainty lexicon significantly reduced the number of expressions used by radiologists in prostate MR reports, without compromising diagnostic accuracy.^[12]^ Structured reporting templates, such as LI-RADS and PI-RADS, can also be regarded as standardized lexicons, and their effectiveness in clinical practice has been well documented.^[13–15]^ However, these systems were designed for a limited number of specific diseases. To broadly enhance communication across radiology reporting, a universally applicable standardized lexicon is required.

Our study had several limitations. The first limitation is the inherent selection bias associated with survey-based data. Second, this was a single-center study with a small sample size. Third, the selection of the referring physicians was not based on a fully systematic or randomized approach. Although efforts were made to ensure a balanced distribution of clinical departments and years of experience, the selection process may have introduced selection bias. Fourth, although the median values for “ari (present)” (100% vs. 100%) and “wa kangae nikui (unlikely)” (10% vs. 10%) were identical between groups, statistically significant differences were observed. This likely reflects a limitation of the Mann–Whitney U test when applied to discontinuous data, where differences in distribution rather than central tendency can yield statistical significance. Such findings may represent statistically significant but clinically negligible differences. Fifth, because the surveys in this study were conducted in Japanese, generalizability to other languages may be limited. Further studies in other languages are required. Sixth, we did not perform a systematic review of our radiology reports to identify the most commonly used modifying terms but relied on solicited opinions from attending radiologists. Finally, our results could be affected by using “metastatic disease” in the survey rather than a more benign diagnosis, such as pneumonia. Furthermore, the anatomical sites associated with “metastatic disease” may have varied among respondents. Depending on their clinical background and experience, some physicians may have envisioned metastases to different organs (e.g., the liver, lungs, and bones), which could potentially influence their interpretation of the disease’s likelihood.

## Conclusion

This study revealed that seven of 15 Japanese expressions describing probability, commonly used in radiology reporting, showed a significant difference in perceived diagnostic probability between radiologists and referring physicians. These findings highlight the need for a standardized lexicon to improve the clarity and consistency of radiology reporting.

## Supporting information

Acknowledgements

## Data Availability

All data produced in the present study are available upon reasonable request to the authors

