## Appendix for "Discrepancies in Interpretation of Japanese Radiology Report Expressions Between Radiologists and Referring Physicians"

**Appendix 1. Copy of the Electronic Survey Distributed to the Radiologists and Referring Physicians (Original)**

**読影レポート表現調査**

**診療科**　　・内科系（放射線科を除く）　　・外科系　　・救急科　　・放射線科　　・その他＿＿＿＿＿

**医師年数（年）**※初期研修期間を含む年数を数字で入力してください　　＿＿＿＿＿

（放射線科医）以下の表現を読影レポートに記載する場合、実際に転移病変である可能性はどの程度と考えますか。最も当てはまる値を選んでください。

（依頼医）以下の表現が読影レポートに記載された場合、実際に転移病変である可能性はどの程度と考えますか。最も当てはまる値を選んでください。

**転移の可能性あり**	・0%　・10%　・20%　・30%　・40%　・50%　・60%　・70%　・80%　・90%　・100%

**s/o 転移**	・0%　・10%　・20%　・30%　・40%　・50%　・60%　・70%　・80%　・90%　・100%

**転移を疑う所見なし**	・0%　・10%　・20%　・30%　・40%　・50%　・60%　・70%　・80%　・90%　・100%

**転移あり**	・0%　・10%　・20%　・30%　・40%　・50%　・60%　・70%　・80%　・90%　・100%

**転移は否定的**	・0%　・10%　・20%　・30%　・40%　・50%　・60%　・70%　・80%　・90%　・100%

**転移疑い**	・0%　・10%　・20%　・30%　・40%　・50%　・60%　・70%　・80%　・90%　・100%

**転移を認める**	・0%　・10%　・20%　・30%　・40%　・50%　・60%　・70%　・80%　・90%　・100%

**転移を否定できない**	・0%　・10%　・20%　・30%　・40%　・50%　・60%　・70%　・80%　・90%　・100%

**c/w 転移**	・0%　・10%　・20%　・30%　・40%　・50%　・60%　・70%　・80%　・90%　・100%

**転移と考える**	・0%　・10%　・20%　・30%　・40%　・50%　・60%　・70%　・80%　・90%　・100%

**転移の可能性は低い**	・0%　・10%　・20%　・30%　・40%　・50%　・60%　・70%　・80%　・90%　・100%

**転移を疑う**	・0%　・10%　・20%　・30%　・40%　・50%　・60%　・70%　・80%　・90%　・100%

**転移として矛盾しない**	・0%　・10%　・20%　・30%　・40%　・50%　・60%　・70%　・80%　・90%　・100%

**転移は指摘できない**	・0%　・10%　・20%　・30%　・40%　・50%　・60%　・70%　・80%　・90%　・100%

**転移は考えにくい**	・0%　・10%　・20%　・30%　・40%　・50%　・60%　・70%　・80%　・90%　・100%

**Appendix 2. Copy of the Electronic Survey Distributed to the Radiologists and Referring Physicians (English Translation)**

**Survey on Radiology Report Terminology**

**Clinical Specialty**

・Internal Medicine (excluding Radiology)	・Surgical Specialties

・Emergency Medicine	・Radiology	・Other: __________

**Years of Clinical Experience** (Please enter the number of years, including postgraduate training.) __________

(For Radiologists) When using the following expressions in a radiology report, what do you consider to be the actual likelihood that the lesion represents metastatic disease? Please select the value that best reflects your judgment.

(For Referring Physicians) When the following expressions appear in a radiology report, what do you consider to be the actual likelihood that the lesion represents metastatic disease? Please select the value that best reflects your judgment.

**Possibly metastatic disease**	・0%　・10%　・20%　・30%　・40%　・50%　・60%　・70%　・80%　・90%　・100%

**s/o metastatic disease**	・0%　・10%　・20%　・30%　・40%　・50%　・60%　・70%　・80%　・90%　・100%

**No findings suggestive of metastatic disease**	・0%　・10%　・20%　・30%　・40%　・50%　・60%　・70%　・80%　・90%　・100%

**Metastatic disease is present**	・0%　・10%　・20%　・30%　・40%　・50%　・60%　・70%　・80%　・90%　・100%

**Negative for metastatic disease**	・0%　・10%　・20%　・30%　・40%　・50%　・60%　・70%　・80%　・90%　・100%

**Suspect of metastatic disease**	・0%　・10%　・20%　・30%　・40%　・50%　・60%　・70%　・80%　・90%　・100%

**Metastatic disease is observed**	・0%　・10%　・20%　・30%　・40%　・50%　・60%　・70%　・80%　・90%　・100%

**Metastatic disease cannot be ruled out**	・0%　・10%　・20%　・30%　・40%　・50%　・60%　・70%　・80%　・90%　・100%

**c/w metastatic disease**	・0%　・10%　・20%　・30%　・40%　・50%　・60%　・70%　・80%　・90%　・100%

**Metastatic disease is considered**	・0%　・10%　・20%　・30%　・40%　・50%　・60%　・70%　・80%　・90%　・100%

**Possibility of metastatic disease is low**	・0%　・10%　・20%　・30%　・40%　・50%　・60%　・70%　・80%　・90%　・100%

**Metastatic disease is suspected**	・0%　・10%　・20%　・30%　・40%　・50%　・60%　・70%　・80%　・90%　・100%

**Not inconsistent with metastatic disease**	・0%　・10%　・20%　・30%　・40%　・50%　・60%　・70%　・80%　・90%　・100%

**Metastatic disease cannot be identified**	・0%　・10%　・20%　・30%　・40%　・50%　・60%　・70%　・80%　・90%　・100%

**Metastatic disease is unlikely**	・0%　・10%　・20%　・30%　・40%　・50%　・60%　・70%　・80%　・90%　・100%
