## Supplementary material for "Discrepancies in Interpretation of Japanese Radiology Report Expressions Between Radiologists and Referring Physicians": Acknowledgements

**Acknowledgments**

The authors thank Munenobu Nogami, Yoshiko Ueno, Akihiro Umeno, Erika Matsushiro, Kentaro Nishiuchi, Daigo Kobayashi, and Naoya Ebisu at Kobe University Graduate School of Medicine for their valuable assistance in recruiting the survey participants. The authors also thank all radiologists and referring physicians who generously contributed their time and insights by participating in the survey.
